# Suicidal behavior during the Covid-19 pandemic: Results from cross-sectional surveys of Brazilian adults from 2020 to 2022

**DOI:** 10.1101/2024.03.06.24303895

**Authors:** André Faro, Dandara Palhano, Luanna dos Santos Silva, Derek Falk

**Affiliations:** Department of Psychology, Federal University of Sergipe, São Cristóvão, Sergipe, Brazil; Department of Population and Quantitative Health Sciences, Case Western Reserve University, Cleveland, Ohio, United States of America

**Author notes:** These authors contributed equally to this work (André Faro, Dandara Palhano). These authors contributed equally to this work (Luanna dos Santos Silva, Derek Falk). Department of Psychology, Federal University of Sergipe, São Cristóvão, Sergipe, Brazil (André Faro).

**Keywords:** Covid-19, suicidal behavior, fear of Covid-19, anxiety, depression

## Abstract

This study aimed to analyze the predictive capacity of fear of Covid-19, anxiety and depressive symptoms, and the variable pandemic year on suicidal behaviors in the years 2020, 2021, and 2022. The total sample consisted of 7,735 adults from all regions of the country and over 1.300 Brazilian cities, with an average age of 34 years. The research instruments used were the Fear of Covid-19 Scale (FCV-19S), the Generalized Anxiety Disorder Scale (GAD-7), the Patient Health Questionnaire Scale (PHQ-9), and a question assessing the absence, presence of suicidal ideation, and suicide planning or attempts. Data analysis was conducted using multinomial logistic regression. The main findings showed that fear of Covid-19 (severe [OR = 0.696]), anxiety (mild [OR = 2.183], moderate [OR = 2.436], and severe [OR = 2.757]), and depression (mild [OR = 2.831], moderate [OR = 4.769], and severe [OR = 10.660]) were statistically significant in relation to suicidal ideation. As for suicide planning and attempts, predictors included year (2022 [OR = 1.297]), fear of Covid-19 (moderate [OR = 0.614] and severe [OR = 0.445]), anxiety (mild [OR = 2.253], moderate [OR = 2.988], and severe [OR = 3.577]), and depression (mild [OR = 3.021], moderate [OR = 8.189], and severe [OR = 40.363]). In conclusion, the effects of the pandemic need to continue to be monitored, as mental health surveillance data are crucial in guiding effective policies aimed at suicidal behavior.

## Introduction

Even in the beginning of 2024, the Covid-19 pandemic continues to result in deaths or significant proportions of sequelae in society [1] [2]. While its numbers have decreased in various parts of the world, such as in China, Portugal, and Ethiopia [2], particularly due to population vaccination [3], it remains - and will likely persist as - a central element in global healthcare [4], including in Brazil [5]. The public health context calls for studies on the repercussions observed in populations since 2020. It is in this direction that the current investigation is proposed: to understand what happened to Brazilian society at a given moment in the years 2020 to 2022 regarding suicidal behavior during the pandemic.

Preventive actions against suicidal behaviors are global imperatives. The WHO [6] began alerting governments and communities about the need to make the prevention of these behaviors a high-priority goal in 2014. Despite these efforts, only 38 of the 138 WHO member countries have a national suicide prevention strategy. Brazil only implemented the National Policy for the Prevention of Self-Harm and Suicide [7] in 2019. This law provided support for a comprehensive and integrated national response to suicidal behaviors. With the onset of the pandemic in 2020, the issue returned to the forefront, given expectations of changes in prevalence throughout the different critical phases of the pandemic [8] [9].

Suicidal behavior encompasses a range of actions, not only the act itself, but also thinking, planning, and attempting suicide [6]. Suicidal ideation involves thoughts about self-destruction, suicidal planning is defined as the formulation of a specific plan to cause one’s own death, and suicide attempt refers to engaging in potentially lethal behaviors with the intention to die. In turn, death by suicide is defined as intentionally taking one’s own life [10]. Epidemiological data help to understand the importance of prevention on the global public health agenda. An estimated 700.000 people die by suicide annually worldwide [11]. Evidence regarding the effects of the pandemic on suicidal behaviors remains relatively scarce and with contradictory data [12] [9], highlighting the need for further research. What is known is that environmental stressors play a significant role in the development of psychological problems. In this case, a pandemic can be considered a powerful disruptive event capable of eliciting stressors that can increase levels of psychological distress for many individuals [13].

Fear has been, and still is, among the most prevalent psychological responses to pandemic events [14]. Fear is defined as an adaptive emotion activated by the assessment of the threat level of a real or imaginary stimulus, but in excess, it can become harmful [15]. Recent studies reveal the role played by the fear of Covid-19, with evidence of both its protective action in motivating good adherence to safety measures [15] [16] and the detrimental outcomes resulting from the persistence and intensification of this emotion [17] [18]. The key to understanding lies in the modulation of fear in relation to the threat; how realistic an individual’s perception of fear is in the face of the actual harm that may result from its encounter. Even with exposure, it is important to understand how capable, skilled, or confident a person feels in dealing with the fear-inducing event. For example, a higher level of fear of Covid-19 has been associated with a higher likelihood of exhibiting suicidal behavior in Taiwan and Spain [19] [20], as well as symptoms of anxiety and depression [17] as evidenced in a systematic review conducted in over 30 countries.

The development or worsening of psychopathological symptoms can influence one’s ability to cope with the adversities caused by the pandemic. Psychological issues such as anxiety and depression are among the primary predictors of suicidal behaviors during the pandemic years and require surveillance due to the expected increases in complaints and demands for assistance services for these conditions [21] [9]. An international meta-analysis observed significantly higher levels of depression and anxiety during the pandemic compared to the pre-pandemic period [22]. It is estimated that since the onset of the health crisis caused by the novel coronavirus, the prevalence of anxiety and depression has increased by 25% worldwide [23]. In Brazil, data collected during the height of the pandemic between May and July 2020 revealed a prevalence of nearly 82% for anxiety and 68% for depression [24].

The psychological repercussions resulting from the various stressors imposed by the pandemic in its different stages have been shown to be increasing and long-lasting [22] [25]. Just as the economic, commercial, and political effects, the psychological effects of this crisis will persist beyond its duration. Investigating the impact of the pandemic on suicidal behaviors, as well as assessing the best strategies to reduce this harm, needs to be prioritized [26]. In other words, it is a macro-level phenomenon in terms of public health, encompassing all aspects of well-being. Among them, mental health is crucial for understanding how the population perceived, coped with, and suffered the consequences of this period.

Therefore, fear of Covid-19, anxiety, and depression can be understood as potential underlying factors for suicidal behaviors. Investigating the relationship between these variables can help depict risk stratification and assist in directing interventions to groups at higher vulnerability. The objective of this study was to analyze the explanatory capacity of levels of fear of Covid-19, anxious and depressive symptoms, as well as the year of the pandemic, on suicidal behaviors in the years 2020, 2021, and 2022, using a sample of Brazilian adults. Additionally, the distribution of the levels of these variables over this period was assessed.

## Materials and methods

### Participants

The final sample consisted of three cross-sectional surveys in the years 2020 (n = 4.793), 2021 (n = 1.476), and 2022 (n = 1.466), totaling 7.735 adults. The average age was 34.0 years (SD = 12.89; Min = 18 and Max = 70), with the majority being females (87.6%; n = 6.778), having a higher level of education (83.8%; n = 6.485), and identifying as white individuals (55.0%; n = 4.252). Participants were residents of approximately 1,300 Brazilian cities. The Northeast region was predominant (44.4%; n = 3.433), followed by the Southeast (34.2%; n = 2.646), South (12.3%; n = 950), Central-West (5.9%; n = 454), and North (3.3%; n = 252). Table 1 presents the distribution of all the collected sociodemographic data.

**Table 1.** Sociodemographic profile (Brazil, n=7,735).

| Variables | Total (n = 7,735) | 2020 (n = 4,793) | 2021 (n = 10,476) | 2022 (n = 1,466) |
| --- | --- | --- | --- | --- |
| Age M(SD) | 34.0 (12.89) | 31.2 (11.78) | 39.1 (13.35) | 37.7 (13.37) |
|  | *F%(n)* | *F%(n)* | *F%(n)* | *F%(n)* |
| Sex |  |  |  |  |
| Male | 12.4 (957) | 13.0 (625) | 10.0 (148) | 12.6 (184) |
| Female | 87.6 (6778) | 87.0 (4168) | 90.0 (1328) | 87.4 (1282) |
| Skin color |  |  |  |  |
| White | 55.0 (4252) | 52.3 (2505) | 67.5 (996) | 51.2 (751) |
| Black | 10.0 (774) | 11.6 (555) | 6.8 (100) | 8.1 (119) |
| Parda | 35.0 (2709) | 36.2 (1733) | 25.7 (380) | 40.7 (596) |
| Educational level |  |  |  |  |
| Up to elementary school | 0.7 (56) | 0.4 (21) | 0.4 (6) | 2.0 (29) |
| High school | 15.4 (1194) | 23.2 (1111) | 1.2 (17) | 4.5 (66) |
| Higher school | 83.8 (6485) | 76.4 (3661) | 98.4 (1453) | 93.5 (1371) |

### Instruments

The Fear of Covid-19 Scale (FCV-19S) [14] is a unidimensional scale containing seven items using a Likert scale from 1 (strongly disagree) to 5 (strongly agree). The total score is obtained by summing the items, ranging from 7 to 35 points. In the adaptation study for Brazilian Portuguese, satisfactory psychometric results were found [27], and the level of fear was classified into three categories: 7 to 19 points as mild fear, 20 to 26 points as moderate, and 27 points and above as severe. In this study, the Cronbach’s alpha (α) was 0.87, and McDonald’s omega (*ω*) was 0.88.

The Generalized Anxiety Disorder Scale - 7 (GAD-7) [28] consists of seven items, which are answered on a scale from 0 (not at all) to 3 (nearly every day) points. The total score ranges from 0 to 21, and the level of anxiety is classified as minimal (0-4 points), mild (5-9 points), moderate (10-14 points), and severe (15-21 points). The psychometric properties and validity evidence for this scale in Brazil are considered satisfactory [29]. In this study, satisfactory internal consistence indices were found (α = 0.89; *ω* = 0.89).

The Patient Health Questionnaire (PHQ-9) [30] consists of 9 items using a scale from 0 (not at all) to 3 (nearly every day). The total score ranges from a minimum of 0 to a maximum of 27. The stratification of scores proposes four categories: 0-4 as minimal, 5-9 as mild, 10-14 as moderate, and 15-27 as severe. The scale has demonstrated satisfactory psychometric characteristics in the Brazilian population [31]. In this study, the Cronbach’s alpha and McDonald’s omega were both 0.90.

The sociodemographic questionnaire includes questions about the participant’s sex (female or male), age (in years), skin color (white, black, parda), education level (up to elementary school, high school, or higher school), and city of residence. Participants were asked if at any point during the pandemic they had thoughts, plans, and/or attempts to end their own life, with the following response options: (0) Never, (1) I had only a brief passing thought, (2) I had a plan to end my life at least once, but I didn’t attempt it, (3) I attempted to end my life, but didn’t really want to die, and (4) I attempted to end my life and actually wanted to die.

### Procedures and Ethical Aspects

This research was approved by the National Commission of Research Ethics (CONEP, registration: 3.955.180). Participant recruitment was conducted through convenience sampling, using social media platforms such as Facebook and Instagram. Only individuals aged 18 or older and those who agreed to the Informed Consent Form, which was presented at the beginning of the online questionnaire, were eligible for the study. Data collection took place during three different periods of the pandemic in Brazil: June 2020, March 2021, and May 2022.

### Data Analyses

The Jamovi software (version 2.2.5) was used for data analyses. A hierarchical multinomial logistic regression was conducted with suicidal behavior as the dependent variable. This variable was recoded into the following groups: (1) absence of suicidal behavior; (2) presence of suicidal ideation; (3) suicide planning and/or attempt. The explanatory variables included the year of data collection (2020, 2021, 2022), and the levels of fear of Covid-19 (7-9 mild, 20-26 moderate, 27-35 severe), anxiety (0-4 no symptoms, 5-9 mild, 10-14 moderate, 15-21 severe), and depression (0-9 no symptoms, 10-14 mild, 15-19 moderate, 20-27 severe), added in this order to the model. Model fit evaluation criteria included the initial and final -2 log likelihood, the significance criteria (p <0.05) of the odds ratios (OR), the model’s predictive ability (correctly predicted cases, with values desired to be above 50%), the X^2^ (expected to be significant at p <0.05), the Pearson Goodness of Fit X^2^ (expected value to be non-significant), and the Pseudo R-Square index (higher values indicate better fit). Odds ratios (OR) values below 1 were converted using the formula 1/OR for standardized result reporting.

## Results

Over the three-year period of the Covid-19 pandemic, the Fear of Covid-19 Scale (FCV-19S) had an average score of 22.6 (standard deviation [SD] = 6.04), with the lowest average score in 2022 (mean [M] = 20.2; SD = 6.23) and the highest in 2021 (M = 23.9; SD = 5.59). In terms of severity, severe fear was proportionally higher in 2021 (34.6%, n = 511), as well as moderate fear in 2021 (44.3%, n = 653). The mild level had the highest value in 2022 (45.4%, n = 666). In total, there was a predominance of moderate fear (42.2%; n = 3.262). The GAD-7 had an overall average of 11.4 points (SD = 5.78), with the highest value in 2022 (M = 12.0; SD = 5.69). Only 12.5% (n = 968) did not exhibit significant anxiety symptoms over the three years surveyed. The proportion of individuals classified with severe anxiety was highest in 2022 (39.0%; n = 572). In contrast, the number of participants at the mild level decreased from 2020 to 2022, dropping from 29.7% (n = 1.423) to 24.0% (n = 352).

The PHQ-9 had an overall average score of 13.4 (SD = 7.44). In the total sample, 34.5% (n = 2.671) did not exhibit significant depression symptoms, with the largest portion reporting severe scores (25.4%; n = 1.966). The highest average score was observed in 2022 (M = 13.9; SD = 7.61). The highest concentration of people classified in the severe level occurred in 2022 (29.7%; n = 435), the same year with the lowest value in the category without depression (32.8%; n = 481). The presence of suicidal ideation, planning, or attempt occurred in 32.1% of the total sample (n = 2.485).

Specifically, suicidal ideation fluctuated slightly, hovering around 20% from 2020 to 2022. However, suicide planning and/or attempts nearly doubled in value, going from almost 10% in 2020 to almost 17% in 2022. Additional details and values obtained in the samples are detailed in Table 2.

**Table 2.** Distribution of Suicidal Behavior Categories and PHQ, GAD, and FCV-19S scales.

| Variables | Total | 2020 | 2021 | 2022 |
| --- | --- | --- | --- | --- |
|  | *M(SD)* | *M(SD)* | *M(SD)* | *M(SD)* |
| FCV-19S score | 22.6 (6.04) | 22.9 (5.92) | 23.9 (5.59) | 20.1 (6.23) |
| GAD score | 11.4 (5.78) | 11.2 (5.83) | 11.9 (5.63) | 12.0 (5.69) |
| PHQ score | 13.4 (7.44) | 13.2 (7.39) | 13.3 (7.40) | 13.9 (7.61) |
|  | *F%(n)* | *F%(n)* | *F%(n)* | *F%(n)* |
| <u>Fear of Covid-19</u> |  |  |  |  |
| Mild | 30.3 (2344) | 28.5 (1366) | 21.1 (312) | 45.4 (666) |
| Moderate | 42.2 (3262) | 42.7 (2045) | 44.2 (653) | 38.5 (564) |
| Severe | 27.5 (2129) | 28.8 (1382) | 34.6 (511) | 16.1 (236) |
| <u>Anxiety symptoms</u> |  |  |  |  |
| No | 12.5 (968) | 13.9 (664) | 9.9 (146) | 10.8 (158) |
| Mild | 28.1 (2171) | 29.7 (1423) | 26.8 (396) | 24.0 (352) |
| Moderate | 24.2 (1873) | 23.1 (1109) | 25.7 (380) | 26.2 (384) |
| Severe | 53.2 (2723) | 33.3 (1597) | 37.5 (554) | 39.0 (572) |
| <u>Depression symptoms</u> |  |  |  |  |
| No | 34.5 (2671) | 35.4 (1696) | 33.5 (494) | 32.8 (481) |
| Mild | 19.7 (1523) | 19.3 (927) | 21.7 (321) | 18.8 (275) |
| Moderate | 20.4 (1575) | 20.9 (1000) | 20.3 (300) | 18.8 (275) |
| Severe | 25.4 (1966) | 24.4 (1170) | 24.5 (361) | 29.7 (435) |
| <u>During the quarantine/social isolation, did you ever think about killing yourself or try to kill yourself?</u> |  |  |  |  |
| Never | 67.9 (5250) | 69.8 (3344) | 67.6 (998) | 61.9 (908) |
| I had only a brief passing thought | 20.8 (1607) | 20.5 (984) | 21.0 (310) | 21.3 (312) |
| I made a plan to kill myself at least once, but I didn't try | 9.6 (744) | 8.7 (415) | 9.6 (142) | 12.8 (187) |
| I tried to kill myself, but I didn't want to die | 0.8 (62) | 0.6 (30) | 0.5 (8) | 1.6 (24) |
| I tried to kill myself, and I really hoped to die | 0.9 (73) | 0.4 (20) | 1.2 (18) | 2.4 (35) |
| <u>Suicidal behavior</u> |  |  |  |  |
| No | 67.9 (5250) | 69.8 (3344) | 67.6 (998) | 61.9 (908) |
| Ideation | 20.8 (1606) | 20.5 (984) | 21.0 (310) | 21.3 (312) |
| Planning or attempt | 11.4 (879) | 9.7 (465) | 11.4 (168) | 16.8 (246) |

### Multinomial Logistic Regression

The final model (Table 3) demonstrated an acceptable solution at all levels (p <0.001). It achieved a variance explained of 20.7% (Nagelkerke’s R^2^ = 0.207), and the correct predictive capacity had a total value of 34%. All the variables included remained as statistically significant in the final model.

**Table 3.**
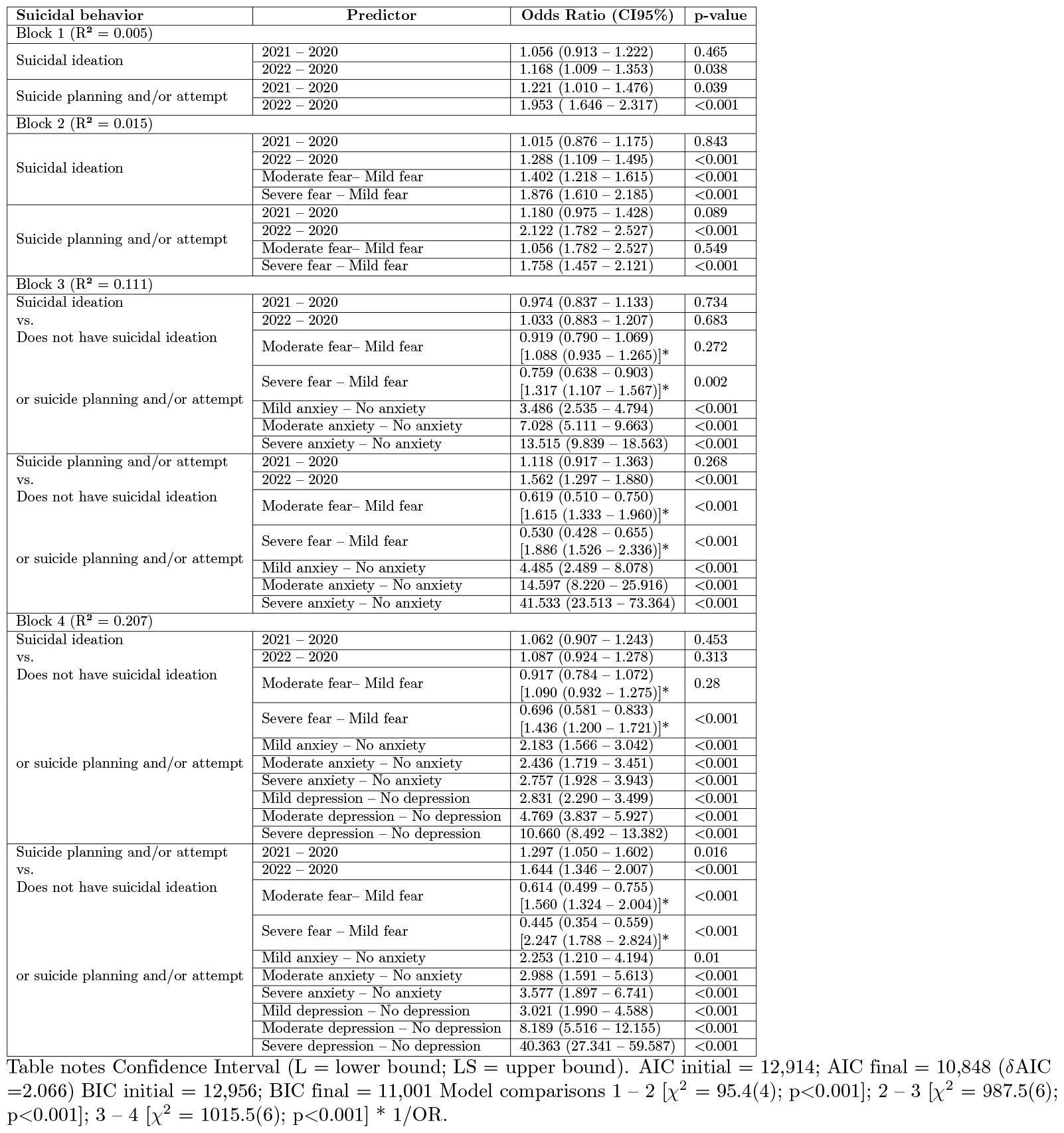
Hierarchical Multinomial Multiple Regression - Reference group “did not present suicidal ideation, planning, or attempt,” (Brazil, n = 7,736).

#### Block 1: Year

The first block, which included the year of data collection, demonstrated an adequate fit indicator [*χ*^2^ = 56.4 (4); p <0.001]. In 2022, there was an additive impact, increasing the odds of being in the group with suicidal ideation by 16.8% (OR = 1.168; p = 0.038) and nearly doubling the odds of suicide planning and/or attempt (OR = 1.953; p<0.001) compared to the year 2020 (OR = 1.221; p <0.039). There was no statistical significance for the contrast between the years 2021 and 2020 for suicidal ideation (p ¿ 0.05).

#### Block 2: Fear of Covid-19

The second block also showed an adequate fit indicator for the FCV-19S scale [*χ*^2^ = 95.4 (4); p <0.001]. Individuals with severe fear (OR = 1.876; p <0.001) or moderate fear (OR = 1.402; p <0.001) had a higher chance of being in the group with suicidal ideation compared to those with mild fear. In this block, the year variable remained significant for 2022, and there was an increase in the odds for both suicidal ideation and suicide planning and/or attempts.

#### Block 3: Anxiety

In this block, there were also appropriate fit indicators for the GAD-7 scale [*χ*^2^ = 987.5 (6); p <0.001]. However, in this block, the direction of the FCV-19S had reversed compared to the second block. The severe fear group had a lower chance of being in the groups with suicidal ideation and suicide planning and/or attempts when compared to mild fear by 31.7% (1/OR = 1.317; p = 0.002) and 88.6% (1/OR = 1.886; p <0.001), respectively. In this block, the year variable was only significant for the year 2022 regarding suicide planning and/or attempts, showing a reduction in odds compared to the second block, by almost two times to 56.2%, when comparing the years 2022 and 2020 (OR = 1.562; p <0.001).

The anxiety variable increased the odds from three and a half times at the mild anxiety level (OR = 3.486; p <0.001) to over 13 times at the severe anxiety level (OR = 13.515; p <0.001) for suicidal ideation. For suicide planning and/or attempts, this range was from four and a half times at the mild anxiety level (OR = 4.485; p <0.001) to over 41 times at the severe anxiety level (OR = 41.533; p <0.001), compared to the group without anxiety.

#### Block 4: Depression

The fit indicators were significant for the PHQ-9 scale [*χ*^2^ = 1015.5 (6); p <0.001]. The year 2022 showed a significant result of 64.4% for suicide planning and/or attempts (OR = 1.644; p <0.001) compared to the year 2020. The fear variable remained significant in the following models. The more severe the fear, the lower the chance of being among those with suicidal ideation, with values of 43.6% (1/OR = 1.436; p <0.001) for severe fear compared to mild fear. The same pattern was observed for suicide planning and suicidal ideation, with the values of 1/OR increasing from 1.886 to over two times (1/OR = 2.247; p <0.001) for severe fear compared to mild fear. Moderate fear remained stable in terms of values from the third to the fourth block.

In the fourth block, it was observed that the adjusted odds of the anxiety variable decreased compared to the third block, specifically for suicidal ideation. The results ranged from two times (OR = 2.183; p <0.001) to approximately three times (OR = 2.757; p <0.001). For suicide planning and/or attempts, the behavior of the variable was the same, with results between two times (OR = 2.253; p <0.001) and over three times (OR = 3.577; p <0.001).

With the inclusion of the depression variable in this block and the adjustment of odds for suicidal ideation, the probabilities ranged from almost three times (OR = 2.831; p <0.001) to over ten times (OR = 10.660; p <0.001). The same pattern occurred when considering the suicide planning and/or attempts group, with the odds increasing from three times (OR = 3.021; p <0.001) to over 40 times (OR = 40.363; p<0.001) when compared to the group without depression.

In summary, in the first block, the additive impact of the year 2022 compared to 2020 was significant for suicidal behaviors. In the second block, the exposure factor of fear of Covid-19 was noteworthy, as the group of people with severe fear exhibited lower chances of displaying suicidal behaviors than those with mild fear. In the third block, with the inclusion of anxiety, the fear of Covid-19 reversed the direction of the relationship and became a protective factor for these behaviors. Moreover, the presence of anxiety in this block increased the chances of displaying suicide planning and/or attempts from over four times to 41 times compared to the group without anxiety.

Finally, in the fourth block, the odds of anxiety were adjusted compared to the previous block. Considering the suicide planning and/or attempts group, the presence of depression increased the odds from three times to over 40 times compared to the group without depression.

Evaluations of hierarchical multinomial logistic analyses and various indicators (VIF, tolerance, residuals, and the discrepancy of OR in relation to their confidence intervals) did not indicate unsatisfactory quality of the findings. Fig1 1 represents the last block of analysis, containing the adjusted ORs and their respective confidence intervals.

**Fig 1.**
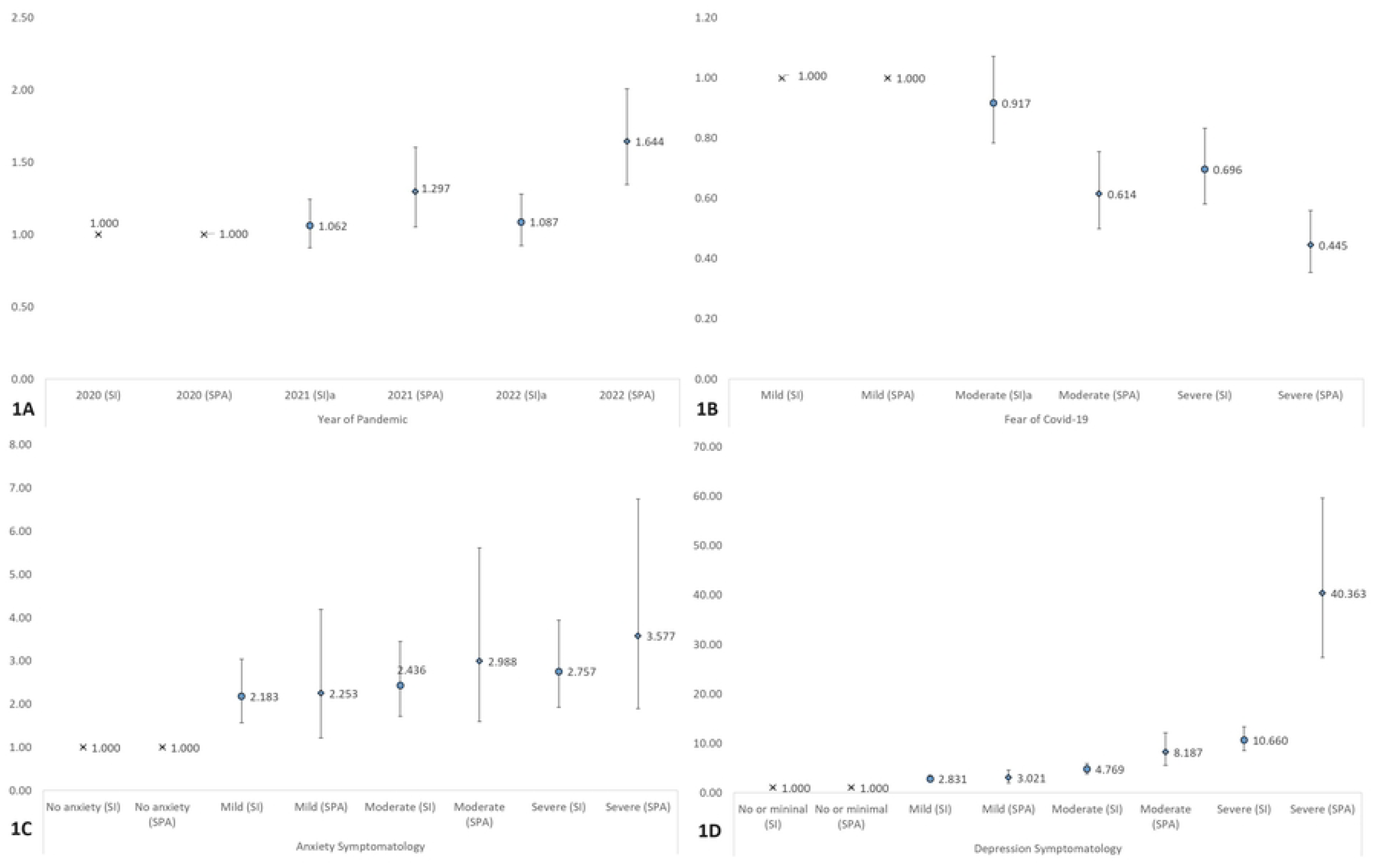
Odds Ratio and Confidence Intervals of the Fourth Block (last model) - Multinomial Logistic Regression for Suicidal Behavior (Brazil, n = 7,736). Notes. Graphical representations of OR and 95% CI in block 4 by variable. 1A: Year of Pandemic. 1B: Fear of Covid-19. 1C: Anxiety Symptomatology. 1D: Depression Symptomatology. a: statistically non-significant relationship.All others were statistically significant.

## Discussion

This study aimed to analyze the predictive capacity of fear of Covid-19, anxiety, and depression on suicidal behaviors in the years 2020, 2021, and 2022. The distribution of these variables over this period were also described. The average level FCV-19S (M = 22.6; SD = 6.04) over the three years of data collection was higher than values found in other countries. For example, in a meta-analysis of studies conducted in 36 countries from December 2019 to June 2021, the average fear of Covid-19 was 13.11 [17], nearly half of the value recorded in this study.

The highest FCV-19S scores occurred in 2021, which seems to reflect the fact that this year was also the period with the highest severity of the disease in Brazil given the number of cases and deaths recorded. Recent data from the Brazilian Institute of Geography and Statistics [32] indicated that 2021 had the highest death rate in the entire historical series, which began in 1974. The number of deaths was concentrated mainly in the first half of the year. In March 2021, when the data for this study were collected, Brazil had over 300,000 Covid-19 deaths and became the country with the highest daily number of deaths from the new coronavirus [33]. The high mortality during this period was attributed to the emergence of new variants of the SARS-CoV-2 virus and the collapse of the healthcare system in various regions of the country [33]. In contrast, participants reported feeling less threatened by the pandemic in 2022, with lower levels of fear of Covid-19. This was the year when Brazil made progress in vaccination, and a decrease in the number of deaths was observed [34]. Despite being slow and uneven, the large-scale vaccine coverage allowed for the relaxation of restriction measures and a return to social interaction [35] [36], possibly leading to changes in the perception of the pandemic’s risk. This finding suggests a potential trend of decreasing fear of Covid-19, as Brazil continues to report a reduction in cases and deaths caused by the coronavirus [37].

The scores of GAD-7 and PHQ-9 increased over the years, with 2022 showing the highest average and the largest number of people in the severe classification category. Most of the sample exhibited significant symptoms of anxiety (87.5%, n = 6776) and depression (65.4%, n = 5069) during the research period. The characteristic pandemic scenario, marked by uncertainties, the imposition of public health measures, illness, and the loss of loved ones, had a severe impact on mental health [38]. Overall, countries in Latin America showed a higher prevalence of depressive and anxious symptoms during the pandemic compared to other regions around the world [39] [40]. This contrast could be attributed to factors such as the severity of the pandemic’s progression in the area, mortality rates, and the ability to adopt coping measures. Additionally, political factors, such as trust in the healthcare system and government, as well as the impact of conflicting messages from authorities, may have also contributed to the identified psychological distress [39] [40].

We found that mental health of the studied population followed a trajectory of worsening, with an increase in the average levels of anxiety and depression from 2020 to 2022, going from 11.2 to 12.0 and 13.2 to 13.9, respectively. After the initial stages of the pandemic, some individuals may have experienced the strain caused by the exhaustion of psychological coping resources, without being able to fully recover their normal functioning [41] [42]. The increase in levels of anxiety and depression observed, especially in 2022, appears to be based on the perspective of an excess of adaptive strain in the context of Covid-19, leading to a vulnerability of mental health [43]. Without time to rebuild the capacity to manage adversity during this period and in the face of the numerous repercussions of the pandemic, the increase in pandemic stress levels was a constant, and perhaps 2022 reflects a peak in this process, at least up to that point.

As with the course of anxiety and depression symptoms, suicidal behavior showed an increase from 2020 to 2022. The growth in the planning and/or suicide attempt category was particularly striking, going from 9.7% to 16.8%. These findings are in line with the results of a meta-analysis that included 45 studies conducted between 2020 and 2022 and identified an upward trend in suicide attempts during the Covid-19 pandemic [9]. In the early stages of the pandemic, it appears that it did not have an immediate impact on suicidal behavior. The collective trauma perception may have normalized the experience of distress and mitigated the suicide risk along with a focus on the hope of survival [26]. However, considering the evolution of the pandemic, the perspective on suicidal behavior may change. For example, a study in Japan recorded a decline in suicide deaths in the early stages of the pandemic, followed by a significant increase from July 2020 onwards [44], indicating possible variations in this behavior over time.

### Predictive variables of the suicidal behavior

When assessing the explanatory power of the pandemic year on suicidal behaviors, we found that in 2022, compared to 2020, the chance of suicidal ideation increased by 16.8%, and the chance of planning and/or attempting suicide nearly doubled. Previous studies had already pointed out the influence of various factors in the increased occurrence of suicidal behaviors, indicating the long-term effects of the Covid-19 pandemic, which would cause a proportional increase in suicide rates during and after the pandemic scenario [45] [46]. The findings of this study align with the existing literature, suggesting that the progression of the pandemic from 2020 to 2022, characterized in Brazil as a growing public health concern [47] [48], seems to have been an explanatory factor for the increased suicidal behavior.

Another finding in this study indicated that a severe or moderate level of fear of Covid-19, when added alongside the pandemic year in the logistic regression model, increased the probability of being in the group with suicidal ideation. Initially, the data found in this study confirmed the literature by suggesting that a greater fear of Covid-19 appears to be associated with suicidal behaviors [19] [20]. Other studies found that fear can have detrimental effects when it persists and intensifies [17] [49] [18], as is the case with the Covid-19 pandemic, which resulted in prolonged exposure to the fear of infection and the presentation of severe forms of the disease. This was particularly impactful in Brazil due to the national policies in place that affected vaccine procurement and even the population’s adherence to vaccination campaigns [48].

When the anxiety variable was included in the model in the third block, the relationship between the fear of Covid-19 and suicidal behavior reversed. In other words, the indication of wanting to take one’s own life (whether ideation or planning and/or attempting) became more pronounced in situations of high anxiety, not high fear of Covid-19. Therefore, fear turned into a protective factor when the effect of anxious symptoms was controlled for in the model, which aligns with another perspective on the analysis of fear of Covid-19 during the pandemic [15] [16]. In terms of logistic regression analysis, this indicates that the anxiety variable had a suppressing effect on the fear of Covid-19 variable [50]. In this case, the second predictor (anxiety) had an indirect effect on the outcome, compensating for the error in the first predictor (fear of Covid-19) and acting as a suppressor. It’s worth noting that the suppressor effect was not observed in the year variable, which maintained the logic of its explanatory capacity. In summary, the results of this research highlighted the need to understand concurrent effects between the two phenomena – fear of Covid-19 and anxious symptomatology – in explaining the occurrence of suicidal behavior during the pandemic.

The findings showed a strong association between anxiety and suicidal ideation, with a risk increase of up to ten times for individuals with severe anxiety. For planning and/or attempting suicide, the association was even higher, reaching more than 40 times among those with severe anxiety, compared to those without anxiety. The high likelihood of suicidal behaviors occurring in individuals with varying levels of anxiety severity is particularly concerning, especially when considering that an estimated one in three adults lived with anxiety during the COVID-19 pandemic worldwide [51]. This underscores the critical importance of addressing mental health issues and providing adequate support for individuals experiencing anxiety and its severe forms, as it relates to suicide risk.

The inclusion of the PHQ-9 in the model was significant and had an impact on reducing the explanatory power of the year, fear of COVID-19, and anxiety. While the relationships of fear and anxiety with the pandemic remained, it was evident that depression accounted for the largest portion of explaining suicidal behaviors during the pandemic. For suicidal ideation, the likelihood of occurrence ranged from nearly three times to more than ten times, depending on the severity level of depressive symptoms. In the group reporting planning and/or attempting suicide, the likelihood increased from three times to more than 40 times when compared to the group without depression. Such advertise becomes more relevant when it is expected that the relationship between “Suicidal attempt” and “suicidal ideation”, in relation to the complete suicide, is stronger for the first one [52]. The current results demonstrate more relevance when show a higher discrepancy in the ORs when the analysis are performed within the group who planned or attempted against the own life. This happened especially when the variable is depression was included in the model, which demonstrated a powerful interrelationship [53].

Other studies also indicate that higher levels of depression appear to be related to the emergence of suicidal behaviors during the COVID-19 pandemic [54] [55]. The emergence of depressive symptoms during the pandemic has been highlighted in various studies worldwide [22] [56], and it remains a particularly relevant concern in a potential post-pandemic scenario as it may be understood as a depletion of adaptive resources [57]. Addressing depression and its associated increased risk for suicidal behaviors is a critical aspect of mental health support during and beyond the COVID-19 pandemic.

Mental health during the pandemic has been the focus of various studies globally, whether assessing associated factors or indicating actions that should be taken to mitigate the effects of the health crisis [53] [58] [59]. This study aimed to contribute by providing information on the occurrence of depression and anxiety symptoms, fear of COVID-19, and the pandemic year as predictors of suicidal behavior. The main findings pointed to the strong influence of depressive and anxious symptoms on suicidal behaviors. When these variables were included in the analysis, the explained variance (adjusted R^2^) was 11% and 20.7%, respectively, reaching a total predictive capacity of 34% when combined with other variables. Additionally, it’s worth noting the role of fear as an exposure variable when analyzed in isolation and as a protective variable when anxiety was added to the model, which required special attention in its interpretation.

It’s also important to consider the pandemic year in data collection, although showed relatively low explanatory potential (adjusted R^2^ = 0.5%), contributed to understanding the transition of phases in the pandemic context.

Several limitations of this study need to be highlighted. Despite having a large sample size, this was a convenience and electronic sample based on digital promotion. Therefore, it can be argued that it reached only people with internet access, which may result in less social diversity. However, it’s worth noting that this factor allowed for respondents from over a thousand Brazilian cities across all states in the country.

Additionally, it should be emphasized that this study did not evaluate important variables for the outcome of suicidal behavior, such as age, gender, or economic and social factors, as pointed out in other studies [45] [60]. This was due to the uneven distribution of proportions in this research, which would have made the findings less reliable. It is important for future studies to control for the effect of these variables on the ones studied here to assess their influence on suicidal behaviors.

While COVID-19 is no longer considered a public health emergency as of April 2023 [61], its consequences, especially those related to mental health, continue to demand surveillance [62]. Some of the consequences of the COVID-19 health crisis will only become apparent over several years, requiring continual monitoring the pandemic’s effects on mental health [63]. The findings of this study align with WHO guidelines [64], which advocate for suicide prevention and emphasize that suicidal behavior remains a concern, especially when considering the pandemic’s consequences. In this regard, it is important that at-risk groups receive prioritized behavioral healthcare attention, such as individuals with depressive and anxiety symptoms, which were the primary predictors of suicide planning and attempts in this study. Suicides are preventable, and early detection of vulnerability factors and specific interventions are essential for this purpose.

## Data Availability

Data cannot be shared publicly because of ethical compromises in front of the National Commission of Research Ethics. Data are available under formal request for researchers who meet the criteria for access to confidential data by contacting the corresponding author via e-mail at.

